# Changes in expenditure on vaping in the context of the rise of disposable vapes and impending regulatory changes: a population study in Great Britain, 2021-2025

**DOI:** 10.1101/2025.06.13.25329552

**Authors:** Sarah E. Jackson, Lion Shahab, Jamie Brown

## Abstract

**Background:** The vaping market in Great Britain has shifted rapidly since 2021, driven by the rise of disposable vapes, regulatory announcements, changing consumer behaviours, and market responses.

**Aim:** To examine changes in the amount people who vape spend on vaping products between 2021 and 2025, overall and within key population subgroups.

**Design:** Monthly representative cross-sectional survey.

**Setting:** Great Britain.

**Participants:** 4,072 people aged ≥16 years surveyed between January 2021 and April 2025 who reported current vaping and provided data on expenditure.

**Main outcome measures:** The outcome was self-reported weekly expenditure on vaping products, adjusted for inflation. Monthly time trends in expenditure were modelled using linear regression, overall and by main device type (disposable/reusable), age, occupational social grade (ABC1=more advantaged/C2DE=less advantaged), smoking status (current/former/never), and (among current cigarette smokers) smoking frequency (daily/non-daily). For context, we also modelled changes in the proportion who reported mainly/exclusive using disposable vapes.

**Results:** Overall, average weekly inflation-adjusted expenditure on vaping products increased from £6.13 (95%CI=£5.66-6.63) to £8.39 (£7.79-9.05) – a relative change of +37.0% – with increases in expenditure occurring predominantly among those using disposable vapes (+42.6%). The trend was, however, non-linear. Expenditure first increased steadily between January 2021 and December 2022, peaking at £9.12 (£8.70-9.57) in September 2023, and declined slightly thereafter. This trend mirrored changes in disposable vape use, which rose from 1.0% (0.5-2.0%) in January 2021 to a peak of 39.8% (36.8-42.8%) in April 2023, then fell to 29.7% (24.8-35.1%) by April 2025. The increases and then declines in expenditure were most pronounced among subgroups whose use of disposables increased and then declined the most. For example, expenditure in those who were aged 16 increased by 143.5% to June 2023 followed by a 30.9% decline by April 2025.

**Conclusions:** From 2021 to 2023, inflation-adjusted overall expenditure on vaping products increased substantially among people who vape in Great Britain, particularly during the rapid rise of disposable vaping. Between 2023 to 2025, overall expenditure declined slightly with large declines in subgroups with the greatest switching away from disposable vapes, particularly young people.

An impending ban on disposables in Britain in 2025, which was announced in 2023, may have been responsible for people switching away from disposables, and unintentionally resulted in vaping becoming more affordable among subgroups, such as young people, who predominantly started vaping with disposables.

## Introduction

The market for vaping products in Great Britain has changed rapidly in recent years, particularly with the rise of disposable vapes since 2021.^1–4^ Understanding how much people spend on these products is critical for informing public health policy, assessing the impact of regulatory measures, and guiding ongoing policy discussions about how best to regulate the nicotine market in a way that deters youth uptake while supporting people who smoke in switching to less harmful alternatives.

In 2023, we published an analysis of trends in expenditure on different nicotine products among adults in England between 2018 and 2022.^5^ This showed that inflation-adjusted weekly expenditure on vaping products was stable between 2018 and late 2020, then increased by 31% up to mid-2022, with the average weekly spend reaching £6.30.^5^ At the time, people who mainly used disposable vapes reported the highest expenditure (£8.41 per week, on average, compared with £6.42 and £5.93 among those who mainly used pod and refillable devices, respectively).^5^

Since 2022, the vaping landscape has continued to evolve. The popularity of disposable vapes surged among young people and adults and became the dominant device type,^1,3,4^ prompting government concern over their appeal and environmental impact.^6^ Media attention and policy debates surrounding vaping’s environmental consequences and attractiveness to youth may have also influenced consumer preferences and perceptions of value for money, contributing to shifts in spending behaviour. In January 2024, the UK government announced a forthcoming ban on disposable vapes,^7^ which took effect from 1 June 2025, and signalled broader regulatory changes under the Tobacco and Vapes Bill.^8^ These announcements were followed by a decline in the proportion of vapers using disposables and a plateau in overall vaping prevalence.^9^ Manufacturers have responded by introducing reusable (rechargeable and refillable) devices that mimic the characteristics of disposable vapes and are sold at similar price points,^10^ which may have led to a decline in the average cost of vaping because refills or replacement pods may be cheaper than buying a new device each time.

Further regulatory changes are on the horizon. A new vaping products duty is scheduled to be introduced in October 2026.^11^ This duty, similar to the excise taxes applied to tobacco products, will be levied on vaping products based on their nicotine content, with higher charges applied to products containing higher levels of nicotine. The duty may influence consumer choices, potentially leading people to opt for products with lower nicotine content to avoid higher taxes. This could affect spending patterns, particularly among those who currently use high-nicotine disposables (or ‘disposable-like’ reusables), as they might shift to lower-nicotine products to reduce costs. The introduction of the duty may also drive changes in the product mix, as manufacturers adapt their product lines to cater to cost-conscious consumers.

Understanding vaping expenditure during this period of transition is important for several reasons. First, spending patterns offer insights into how policy announcements and market responses affect consumer behaviour, including shifts between product types. Transitions away from disposable devices – characterised by low up-front costs and higher ongoing expenses – toward reusable products may alter average spending patterns. Second, vaping expenditure has implications for equity and health: cost can be a barrier or incentive for switching from smoking to vaping,^12,13^ and more expensive products may disproportionately affect lower-income users. Third, as policymakers consider further regulation – including pricing and taxation – accurate, up-to-date information on expenditure is needed to forecast the potential impact of fiscal measures and to monitor their effects over time.

This study therefore aimed to update estimates of average weekly expenditure on vaping products in Great Britain and examine how spending has changed from 2021 to 2025 (i.e., since disposable vapes have become popular^2,14^) in the context of evolving product trends and regulatory developments.

## Methods

### Design

Data were drawn from the Smoking Toolkit Study, an ongoing monthly cross-sectional survey of a representative sample of people aged ≥16 years in Great Britain.^15,16^ The study uses a hybrid of random probability and simple quota sampling to select a new sample of approximately 2,450 participants each month. Data are collected through telephone interviews. Comparisons with other national surveys and sales data indicate the survey achieves nationally representative estimates of key sociodemographic and nicotine use variables.^15,17^

### Participants

The present analyses used data from respondents aged ≥16 years who reported current vaping between January 2021 (around six months before disposable vapes started to become popular^2^ and two years before the government’s announcement of new potential vaping policies^18^) and April 2025 (the most recent data on expenditure available at the time of analysis). Data on expenditure were collected in selected waves (January-December 2021; January-April, July, and September-October 2022; January, April, June-July, and October 2023; January, April, July, and October 2024; January and April 2025), so we analysed data from those surveyed in these months. The model used data from these months to estimate expenditure within each month across the entire period. Because the item on expenditure on alternative nicotine products did not differentiate between product groups (see *Measures*), we also restricted the sample to those who reported exclusively vaping (i.e., did not also report using nicotine replacement therapy, heated tobacco products, or nicotine pouches).

### Measures

Vaping status was assessed within several questions asking about use of a range of nicotine products. Participants who reported current smoking were asked ‘Do you regularly use any of the following in situations when you are not allowed to smoke?’ and those who reported cutting down ‘Which, if any, of the following are you currently using to help you cut down the amount you smoke?’; those who currently smoked or who had quit in the past year were asked ‘Can I check, are you using any of the following either to help you stop smoking, to help you cut down or for any other reason at all?’; and those who reported not having smoked in the past year were asked ‘Can I check, are you using any of the following?’. Reporting use of vaping products (‘electronic cigarette’ or ‘Juul’) in response to any of these questions was considered current vaping.

Weekly expenditure on vaping products was assessed with the question: ‘On average about how much per week do you think you spend on using this nicotine replacement product or products?’. This question followed the assessment of current use of vaping products, nicotine replacement therapy, heated tobacco products, and nicotine pouches and referred to the product(s) the participant reported using. We analysed data from participants who reported using only vaping products, to avoid including expenditure on other nicotine products. Participants were asked to only answer the question on expenditure if they were fairly confident that they knew. Responses were given to the nearest pound. We log-transformed expenditure variables for analysis to normalise skewed distributions and reported results as geometric means. Inflation adjustment was calculated using monthly Consumer Prices Index including owner occupiers’ housing costs (CPIH) inflation published by the Office for National Statistics.^19^

The main device type used was assessed with the question: ‘Which of the following do you mainly use…?’ (a) a disposable e-cigarette or vaping device (non-rechargeable); (b) an e-cigarette or vaping device that uses replaceable pre-filled cartridges (rechargeable); (c) an e-cigarette or vaping device with a tank that you refill with liquids (rechargeable); or (d) a modular system that you refill with liquids (you use your own combination of separate devices: batteries, atomizers, etc.). We categorised device types as disposable (response a) vs. reusable (b-d). Participants could also respond that they did not know; these cases were treated as missing. In a sensitivity analysis, we analysed different types of reusable devices separately: pod (response b), tank (c), and mod (d).

Age was analysed as a continuous variable, modelled using restricted cubic splines (see *statistical analysis* section). Socioeconomic position was assessed using the National Readership Survey measure of occupational social grade^20^ and categorised as ABC1 (includes managerial, professional, and upper supervisory occupations) and C2DE (includes manual routine, semi-routine, lower supervisory, state pension, and long-term unemployed).

Smoking status was assessed by asking participants which of the following best applied to them: (a) I smoke cigarettes (including hand-rolled) every day; (b) I smoke cigarettes (including hand-rolled), but not every day; (c) I do not smoke cigarettes at all, but I do smoke tobacco of some kind (e.g. pipe, cigar or shisha); (d) I have stopped smoking completely in the last year; (e) I stopped smoking completely more than a year ago; (f) I have never been a smoker (i.e. smoked for a year or more). Responses were categorised as current smoking (a-c), former smoking (d), and never regular smoking (f). Frequency of cigarette smoking was categorised as daily (a) vs. non-daily (b).

### Statistical analysis

Data were analysed in R v.4.2.2. The Smoking Toolkit Study uses raking to create survey weights that match the sample to the population in Great Britain.^16^ The following analyses were done on weighted data. We excluded participants with missing expenditure data or who reported zero expenditure; missing data on other variables were excluded on a per-analysis basis (see **Table S1** for details). The analyses were not pre-registered and should be considered exploratory.

We used linear regression to model time trends in expenditure on vaping products (dependent variable) from January 2021 to April 2025. We modelled survey month (independent variable) using restricted cubic splines to allow flexible and non-linear relationships with time, increasing the precision and power of results while avoiding arbitrary categorisation. We compared models with three, four, and five knots (sufficient to accurately model trends across years without overfitting) using the Akaike Information Criterion (AIC). The best fitting model was selected as the model with the lowest AIC or the simplest model within two AIC units; three knots provided the best fit (AIC=13624 vs. 13630 and 13635 for models with four and five knots, respectively).

To explore moderation by main device type, age, socioeconomic position, smoking status, and (among those who also smoked cigarettes) frequency of smoking, we repeated the model including the interaction between the moderator of interest and survey month, thus allowing for time trends to differ across subgroups. Each of the interactions was tested in a separate model with time modelled using the same number of knots as in the best-fitting model for the overall trend (this also provided the best fit for each interaction). Age was modelled using restricted cubic splines with three knots (placed at the 1, 50, and 95% percentiles), to allow for a non-linear relationship with expenditure. We displayed estimates for specific ages (16-, 25-, 35-, 45-, 55-, and 65-year-olds) to illustrate how trends differ across ages. Note that the models used to derive these estimates included data from participants of all ages.

For context, we also modelled time trends in the proportion of participants mainly/exclusively using disposable vapes over the study period, overall and by moderating variables. We used a similar approach to the one described above, using logistic regression with disposable use as the dependent variables and survey month (independent variable) modelled non-linearly using restricted cubic splines; four knots provided the best fit (AIC=4198 vs. 4224 and 4200 for models with three and five knots, respectively).

We used predicted estimates from our models to plot (i) the average weekly expenditure on vaping products and (ii) the proportion mainly/exclusively using disposable vapes over the study period – overall and by moderating variables. We reported modelled estimates of expenditure in the first and last months in the time series. Within each calendar year, we also reported unmodelled weighted estimates of the average weekly expenditure on vaping products, and the proportions spending <£5, £5-<£10, £10-<£15, £15-<£20, and ≥£20 per week.

## Results

A total of 70,944 participants were surveyed in eligible waves between January 2021 and April 2025; 5,829 (8.2%) reported current vaping, of whom 5,173 (88.7%) reported no concurrent use of NRT, heated tobacco, or nicotine pouches. Of the latter group, 4,138 (80.0%) reported their weekly expenditure on vaping products and formed the analytic sample. We excluded 993 who responded that they did not know how much they spent and 42 who reported zero expenditure.

The proportion with missing expenditure data was highest among those reporting current smoking (especially daily smoking) and lowest among those reporting former smoking, meaning the analysed sample slightly overrepresented former smokers and underrepresented current smokers (**Table S1**). It was also higher among those with missing data on their main device type and slightly lower among those mainly/exclusively using reusable (vs. disposable) vapes but was similar by age and social grade (**Table S1**).

### Overall trends

Across the entire period, the distribution of weekly expenditure was positively skewed, with most users spending less than £15 per week, though a minority reported substantially higher expenditures (**Figure 1, Table S2**). In 2025, 16.1% reported spending less than £5 a week, 31.3% £5 to <£10, 26.4% £10 to <£15, 10.4% £15 to <£20, and 15.8% £20 or more (**Table S2**).

**Figure 1.**
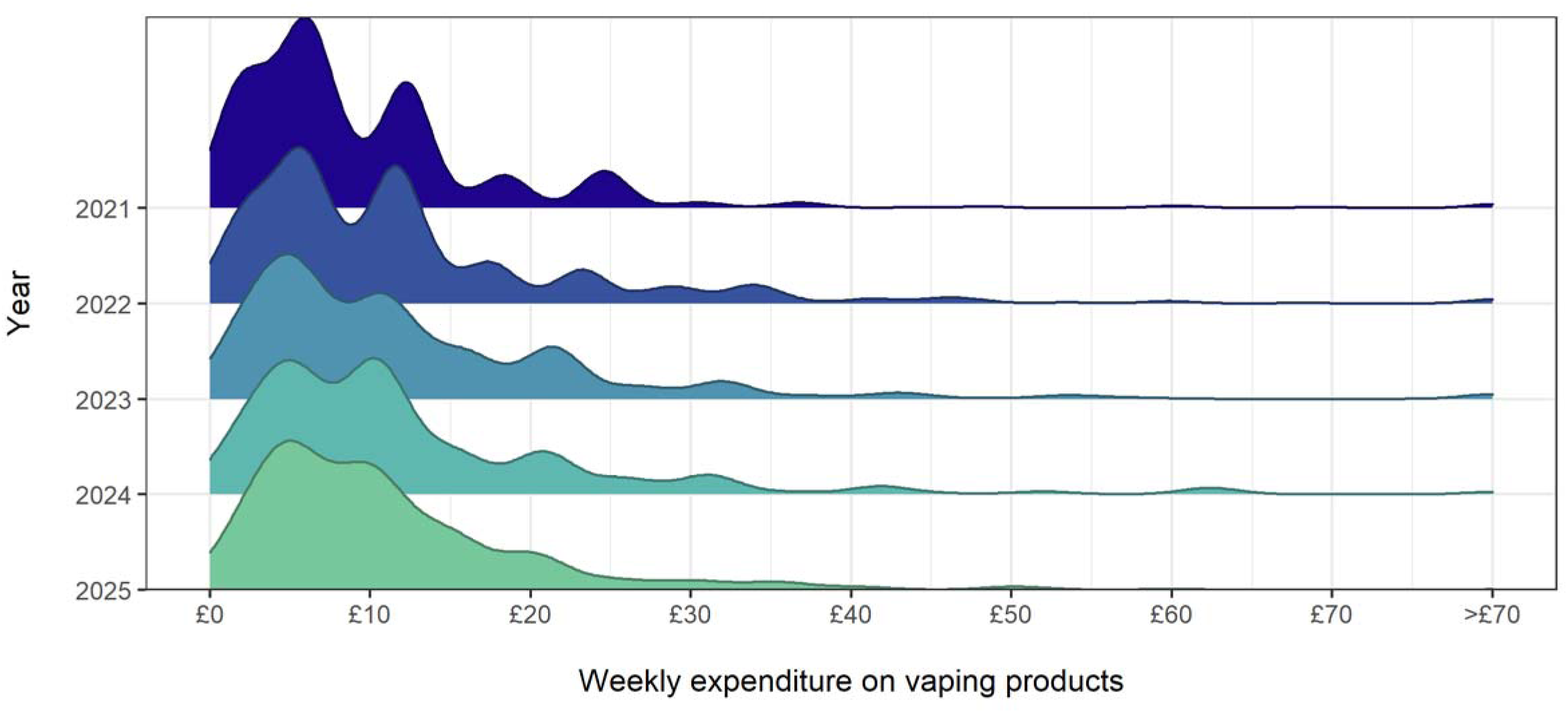
Distribution of weekly inflation-adjusted expenditure on vaping products among people in England who vape, aggregated across the study period (January 2021-April 2025). Unmodelled weighted estimates of geometric mean expenditure within each year are provided in **Table S3**.

From January 2021 to April 2025, the average weekly inflation-adjusted expenditure on vaping products increased by £2.27 [£1.39–2.85]; a relative increase of 37.1% (**Table 1**). This trend was non-linear (**Figure 2A**): average expenditure increased steadily between January 2021 and December 2022 (average +£0.12/month) from £6.13 [95%CI £5.66–6.63] to £8.77 [£8.34–9.32]. Growth slowed between December 2022 and September 2023 (average +£0.04/month), peaking at £9.12 [£8.70–9.57]. There was then an uncertain decline to £8.39 [£7.79–9.05] by April 2025 (average -£0.04/month).

**Figure 2.**
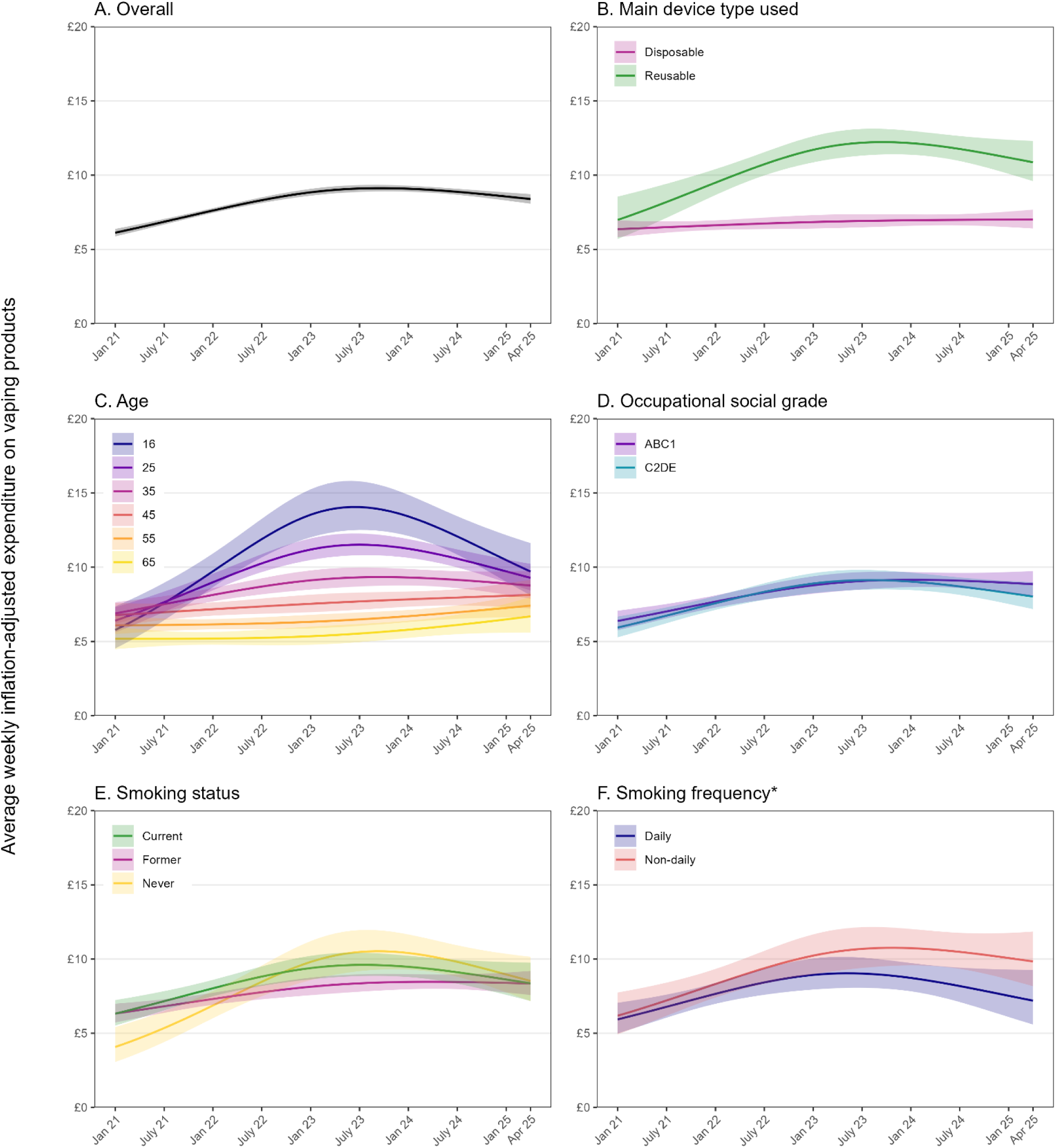
Trends in expenditure on vaping products among people in England who vape, January 2021 to April 2025. Lines represent modelled weighted (geometric) mean inflation-adjusted weekly expenditure (in £) on vaping products by monthly survey wave, modelled non-linearly using restricted cubic splines (three knots), (A) overall and by (B) the main device type used, (C) age, (D) occupational social grade, (E) smoking status, and (F) frequency of smoking among those who also smoke cigarettes. Shaded bands represent 95% confidence intervals.

**Table 1.** Modelled estimates of changes in weekly inflation-adjusted expenditure (in £) on vaping products among people (≥16y) in England who vape.

|  | Mean [95% CI] expenditure (£) |  | Relative change,<br>%<sup>3</sup> |
| --- | --- | --- | --- |
|  | January 2021 | April 2025 |  |
| Overall | 6.13 [5.66–6.63] | 8.39 [7.79–9.05] | +37.0 |
| Main device type |  |  |  |
| Reusable | 6.37 [5.86–6.93] | 7.51 [6.90–8.17] | +17.9 |
| Disposable | 7.97 [5.75–11.06] | 11.37 [9.79–13.20] | +42.6 |
| Age (years)<sup>4</sup> |  |  |  |
| 16 | 5.77 [4.53–7.36] | 9.71 [8.11–11.62] | +68.1 |
| 25 | 6.40 [5.61–7.30] | 9.28 [8.43–10.22] | +45.0 |
| 35 | 6.89 [6.21–7.64] | 8.76 [7.88–9.75] | +27.3 |
| 45 | 6.77 [6.04–7.60] | 8.13 [7.22–9.15] | +20.1 |
| 55 | 6.08 [5.49–6.73] | 7.41 [6.63–8.28] | +21.8 |
| 65 | 5.18 [4.48–5.99] | 6.69 [5.59–7.99] | +29.1 |
| Occupational social grade |  |  |  |
| ABC1 (more advantaged) | 6.38 [5.76–7.07] | 8.86 [8.05–9.74] | +38.8 |
| C2DE (less advantaged) | 5.94 [5.28–6.67] | 8.03 [7.17–8.98] | +35.2 |
| Smoking status |  |  |  |
| Never | 4.07 [3.06–5.41] | 8.52 [7.15–10.15] | +109.3 |
| Former | 6.32 [5.72–6.99] | 8.35 [7.58–9.20] | +32.0 |
| Current | 6.32 [5.52–7.23] | 8.36 [7.18–9.75] | +32.4 |
| Frequency of smoking<sup>5</sup> |  |  |  |
| Daily | 5.93 [4.99–7.05] | 7.19 [5.58–9.26] | +21.2 |
| Non-daily | 6.18 [4.93–7.74] | 9.84 [8.17–11.85] | +59.2 |

CI, confidence interval.
<sup>1</sup> Data are weighted estimates of geometric mean expenditure in the first and last months in the study period, from linear regression with survey month modelled non-linearly using restricted cubic splines (three knots). Unmodelled weighted estimates within each year are provided in **Table S3**.
<sup>2</sup> Absolute percentage point change calculated as expenditure in April 2025 minus expenditure in January 2021 with 95% CIs calculated using bootstrapping (1,000 replications).
<sup>3</sup> Relative percentage change calculated as the difference in expenditure between April 2025 and January 2021 divided by expenditure in January 2021.
<sup>4</sup> Modelled estimates are shown for selected ages to illustrate differences. Note that the model used to derive these estimates included data from participants of all ages, not only those aged 16, 25, 35, 45, 55, or 65 years.

This trend aligned with changes in the proportion who reported mainly/exclusively using disposable vapes (**Figure 3A**), which increased rapidly from 1.0% [0.5–2.0%] in January 2021 to 37.9% [34.7–41.2%] in December 2022, peaked at 39.8% [36.8–42.8%] in April 2023, then fell to 29.7% [24.8–35.1%] by April 2025.

**Figure 3.**
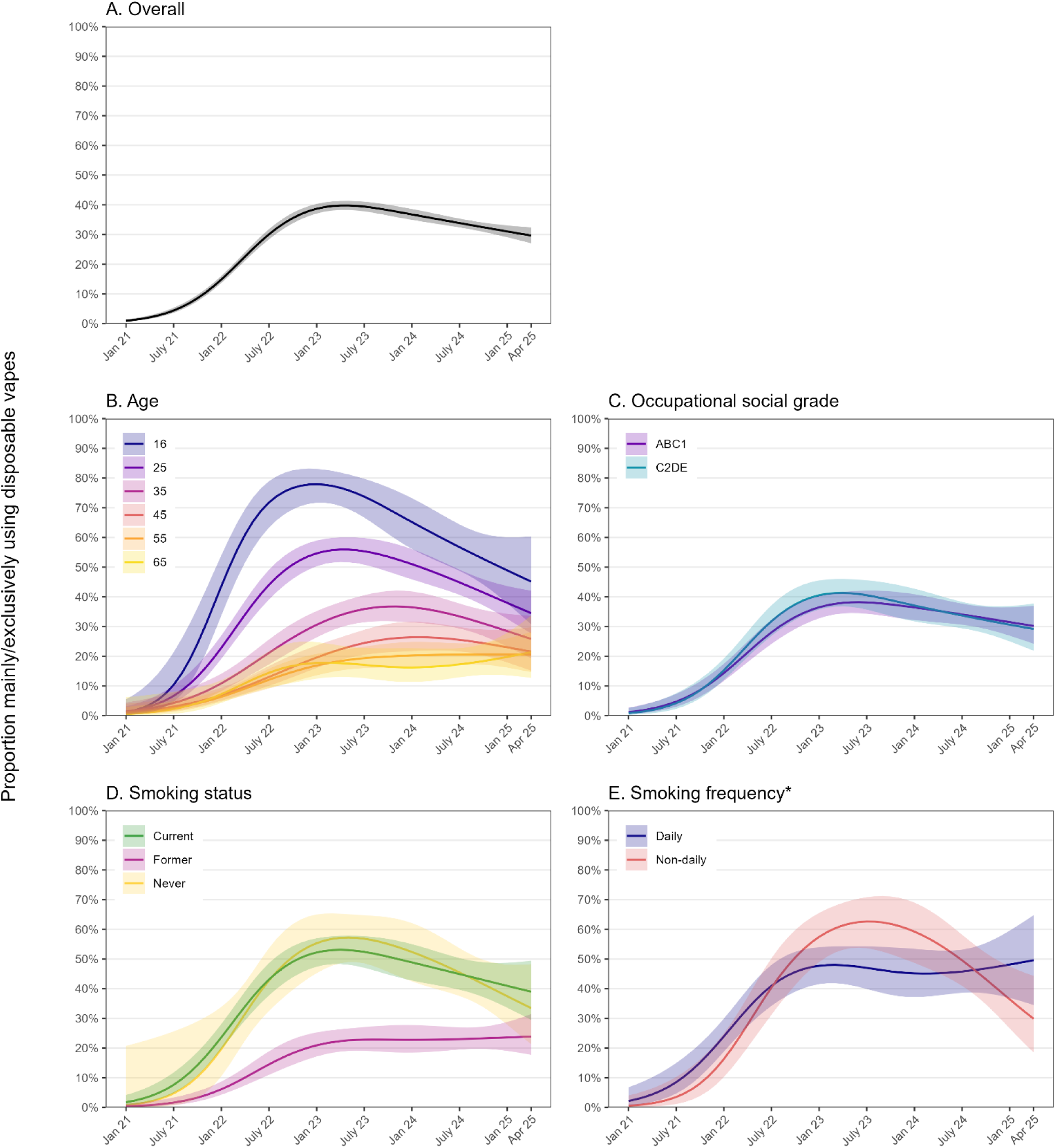
Trends in the proportion mainly/exclusively using disposable vapes among people in England who vape, January 2021 to April 2025. Lines represent modelled weighted prevalence of disposable use by monthly survey wave, modelled non-linearly using restricted cubic splines (four knots), (A) overall and by (B) age, (C) occupational social grade, (D) smoking status, and (E) frequency of smoking among those who also smoke cigarettes. Shaded bands represent 95% confidence intervals.

### Subgroup differences

Although average expenditure on vaping products increased between January 2021 and April 2025 in all population subgroups (**Table 1**), there were some notable subgroup differences in trends (**Figure 2B-F**).

The increase in expenditure occurred predominantly among those using disposable vapes (**Figure 2B**). Among those mainly/exclusively using disposables, changes mirrored the overall trend: average expenditure increased from £7.97 [£5.75–11.06] in January 2021 to £11.50 [£10.58–12.51] in December 2022 (average +£0.15/month), then more slowly between December 2022 and November 2023 (average +£0.05/month), reaching a peak of £12.07 [£11.14–13.09]. There was then an uncertain decline to £11.37 [£9.79–13.20] by April 2025 (average -£0.04/month). In contrast, average expenditure increased modestly among those using reusable devices from £6.37 [£5.86–6.93] in January 2021 to £7.59 [£7.14–8.07] in June 2023 (average +£0.04/month), then remained relatively stable at an average of £7.60 [£7.17–8.06] for the remainder of the period (**Figure 2B**). Sensitivity analyses that distinguished between different types of reusable vapes showed tanks were most commonly used across the entire period (**Figure S1**). Expenditure was relatively stable over time among those mainly/exclusively using tank and mod devices, but among those using pod devices the trend was similar to those using disposables – increasing from £6.51 [£5.01–8.45] in January 2021 to £12.41 [£10.60–14.53] in July 2023, followed by an uncertain decline to £10.00 [£8.01–12.48] by April 2025 (**Figure S1**).

Increases in expenditure were most pronounced among those who were younger (**Figure 2C**) and had never regularly smoked (**Figure 2E**) – groups with higher uptake of disposable vaping (**Figure 3B** and **Figure 3D**, respectively). As these groups increasingly switched away from disposables throughout 2023–2025, their average expenditure on vaping products also decreased the most.

For example, among 16-year-olds, the average weekly expenditure increased from £5.77 [£4.53– 7.26] in January 2021 to a peak of £14.05 [£12.49–15.81] per week in June 2023 (a 143.5% increase), before falling to £9.71 [£8.11–11.62] by April 2025 (a 30.9% decrease from the peak; **Figure 2C**). Correspondingly, disposable use increased from 1.2% [0.3–5.5%] in January 2021 to a high of 77.9% [71.7–83.1%] in January 2023 then falling to 45.2% [30.8–60.4%] by April 2025 (**Figure 3B**). Trends were more modest with increasing age – for example, the proportion using disposables peaked at 55.9% [51.8-60.0%] among 25-year-olds and 36.8% [31.7-42.1%] among 35-year-olds (**Figure 3B**). Among these groups, average expenditure peaked at £11.51 [£10.81-12.27] and £9.34 [£8.76-9.95], respectively – relative increases of 79.8% and 35.6%, compared with January 2021 (**Figure 2C**). Changes in expenditure over time were even less pronounced at older ages, where uptake of disposable vapes was much lower.

Among people who vaped but had never regularly smoked, average weekly expenditure increased from £4.07 [£3.06–5.41] in January 2021 to a peak of £10.53 [£9.28–11.95] in June 2023 (a 158.7% increase) then declined to £8.52 [£7.15–10.15] per week by April 2025 (a 19.1% decrease from the peak), but still remained more than twice as high relative to the start of the period (**Figure 2E**). This group had the lowest starting expenditure in January 2021, but reached parity with those reporting current or former smoking by the end of the study period, among whom changes in expenditure over time were more modest (**Figure 2E**).

Among people who vaped and also smoked cigarettes, expenditure increased more among those who smoked non-daily (**Figure 2F**). Their average weekly expenditure increased from £6.18 [£4.93–7.74] to a peak of £10.76 [£9.55–12.11] in November 2023, then there was a small, uncertain decrease to £9.84 [£8.17–£11.85] – representing a 59.2% overall increase from January 2021. Among those who smoked daily, expenditure increased by just 21.2% (from £5.93 [£4.99– 7.05] to £7.19 [£5.58–9.26]).

Time trends in expenditure and uptake of disposable vapes were similar by social grade (**Figure 2D** and **Figure 3C**).

## Discussion

This study provides an up-to-date picture of trends in average weekly expenditure on vaping products among people aged ≥16 in Great Britain between January 2021 and April 2025, a period marked by rapid market evolution and significant regulatory developments. We observed a substantial increase in inflation-adjusted average weekly spending on vaping products between 2021 and mid-2023, after which spending appeared to stabilise or possibly began to decline. This trend closely tracked the rise and subsequent decline in the use of disposable vapes.

Consistent with previous research,^5^ we found that disposable vapes remained the most expensive option on average. Despite their low upfront cost, their single-use design and frequent replacement contribute to higher ongoing expenses compared to reusable devices. The initial increase in expenditure we observed was concentrated among those using disposable vapes and was most pronounced among those who were younger and had never regularly smoked – groups with higher relative uptake of disposable vaping. Following the UK Government’s announcement in January 2024 of a forthcoming ban on disposable vapes (which came into effect in June 2025),^18^ their use began to fall sharply.^9^ Correspondingly, average expenditure among younger vapers decreased, though it remained higher than before disposable vapes started to become popular. The apparent increase in affordability of vaping among these subgroups since the announcement of the disposables ban, by shifting people towards cheaper vaping products, would be an unintended consequence of the policy.

The high levels of expenditure reported by younger vapers are striking, averaging £9.71 per week among 16-year-olds by April 2025. According to the Office for National Statistics, median weekly earnings for 16-17-year-olds in the UK were approximately £90 in 2024,^21^ meaning vaping expenditure could represent more than 10% of their total income. By comparison, 45-year-olds, spending on average £8.13 per week with median weekly earnings around £700,^21^ spend a much smaller proportion of their income on vaping (∼1.2%). The high expenditure on vaping products by young users raises concerns about the allocation of limited financial resources toward non-essential products and potential knock-on effects (e.g., stress caused by money worries).^22^

Expenditure patterns were broadly consistent across social grades, with similar trends in both vaping spend and uptake of disposables. This suggests that the shifts observed were not limited to a particular socioeconomic group, highlighting the broad reach of disposable vaping products across the population. However, equal uptake does not imply equal impact, as the relative financial burden will differ by income level – an important consideration for future policy evaluation.

Among people who both smoked and vaped, we observed a greater relative increase in vaping expenditure over time among non-daily vs. daily smokers. This may reflect different motivations or consumption patterns, with non-daily smokers likely relying more heavily on vaping as a primary source of nicotine.^23^ However, this difference may also be linked to the higher rate of missing expenditure data among daily vs. non-daily smokers, so should be interpreted with some caution.

Expenditure among those who used reusable vaping products remained comparatively stable over time, reinforcing the idea that recent shifts in average expenditure were driven primarily by changing patterns of disposable vape use. As reusable devices typically involve higher upfront costs but lower ongoing expenses, growing adoption – either voluntarily or in response to regulation – may reduce weekly spending in the longer term. The recent emergence of ‘disposable-like’ reusable products, which replicate the appeal of disposables while offering lower ongoing costs (i.e., cheaper replaceable pods rather than device replacement), may also be contributing to the levelling off in expenditure. In sensitivity analyses that distinguished between different types of reusable vapes, mean expenditure appeared to decrease between 2023 and 2025 among those mainly/exclusively using pod devices. The availability of disposable-like pod devices may lower the financial barrier to continued vaping while also responding to regulatory pressures targeting disposables.

An unintended consequence of the ban on disposable vapes may therefore be to make vaping more affordable for some users. Eventually, this trend will unfold alongside broader fiscal and regulatory developments. The forthcoming duty on vaping products in October 2026,^11^ based on nicotine content, is likely to raise prices for higher-nicotine items and may encourage a shift toward lower-nicotine alternatives.^24^ Additional changes under the Tobacco and Vapes Bill, currently progressing through parliament with cross-party support,^8,25^ are also likely to affect consumer behaviour but these changes may not be implemented before 2027. Continued monitoring will be important to understand how these regulations impact vaping patterns, expenditure, and public health outcomes over time.

This study has several limitations. First, data were self-reported and thus susceptible to recall and social desirability biases. Although participants were asked to report expenditure only if they felt fairly confident in their estimates, they were asked to report weekly averages and inaccuracies may still exist. Second, approximately one-quarter of eligible participants were excluded due to missing expenditure data. If these individuals differed systematically in their spending behaviour (e.g., amount spent, frequency of purchasing, bulk buying, source of purchase) or sociodemographic characteristics, the results may not fully represent the broader population of vapers. Third, our expenditure measure grouped all non-combustible nicotine products. To mitigate this, we restricted the sample to participants who reported exclusively using vaping products, but this means our estimates do not reflect those also using other non-combustible nicotine products (however, this group was relatively small; ∼11% of all adults who vaped). Fourth, subgroup analyses were limited by smaller sample sizes in some groups (e.g., never smokers), reducing precision of estimates, especially at the start and end of the study period. Finally, the observational nature of the data precludes causal inferences about changes in expenditure over time or differences between subgroups. Qualitative research is required to gain further insights.

In conclusion, our findings highlight dynamic shifts in vaping expenditure in Great Britain against a backdrop of regulatory reform and evolving market preferences. Rapidly changing patterns of expenditure, with sharp increases followed by decreases, especially among younger users, and changes in device use underscore the importance of continued surveillance. Policymakers should consider how fiscal measures and product regulations may influence affordability, access, and health equity in vaping behaviours. For a more complete picture, future research should also explore how shifts in product pricing, availability, and regulation impact not only spending but also vaping initiation, motivation to quit vaping, broader nicotine use patterns, and health inequalities.

## Data Availability

Data used in the present study are available upon reasonable request to the authors

## Declarations

### Ethics approval

Ethical approval for the STS was granted originally by the UCL Ethics Committee (ID 0498/001). Participants provide informed consent to take part in the study, and all methods are carried out in accordance with relevant regulations. The data are not collected by UCL and are anonymised when received by UCL.

### Competing interests

JB has received (most recently in 2018) unrestricted research funding from Pfizer and J&J, who manufacture smoking cessation medications. LS has received honoraria for talks, unrestricted research grants and travel expenses to attend meetings and workshops from manufactures of smoking cessation medications (Pfizer; J&J), and has acted as paid reviewer for grant awarding bodies and as a paid consultant for health care companies. All authors declare no financial links with tobacco companies, e-cigarette manufacturers, or their representatives.

## Funding

This work was supported by Cancer Research UK (PRCRPG-Nov21\100002) and the Economic and Social Research Council (ES/Y001044/1). JB is a member of the Behavioural Research UK Leadership Hub which is supported by the Economic and Social Research Council (ES/Y001044/1). For the purpose of Open Access, the author has applied a CC BY public copyright licence to any Author Accepted Manuscript version arising from this submission.

## Notes

### Summary of Updates

Correction to Figure 2B legend

