## Supplementary material for "Changes in expenditure on vaping in the context of the rise of disposable vapes and impending regulatory changes: a population study in Great Britain, 2021-2025": Table S1

**Table S1.** Proportion missing expenditure data and characteristics of analysed and excluded participants

| | | Sample characteristics, column % [95% CI] | | |
|---|---|---|---|---|
| | **Missing expenditure data[1], row % [95% CI]** | **All eligible participants[1]** | **Analysed[2]** | **Excluded due to missing expenditure data[3]** |
| *N* | 5173 | 5173 | 4138 | 1035 |
| Overall | 20.0 [18.9-21.1] | - | - | - |
| Main device type | | | | |
| Reusable | 14.9 [13.7-16.1] | 74.7 [73.5-76.0] | 75.7 [74.3-77.0] | 69.8 [66.4-73.0] |
| Disposable | 19.1 [16.9-21.3] | 25.3 [24.0-26.5] | 24.3 [23.0-25.7] | 30.2 [27.0-33.6] |
| Missing, *n* | 70.9 [66.3-75.4] | 381 | 111 | 270 |
| Age (years) | | | | |
| 16-24 | 19.4 [17.0-21.7] | 20.4 [19.3-21.5] | 20.6 [19.4-21.8] | 19.7 [17.4-22.2] |
| 25-34 | 21.4 [19.1-23.6] | 24.0 [22.9-25.2] | 23.6 [22.3-24.9] | 25.6 [23.0-28.4] |
| 35-44 | 20.3 [17.5-23.0] | 16.3 [15.3-17.4] | 16.3 [15.2-17.4] | 16.5 [14.4-18.9] |
| 45-54 | 17.1 [14.5-19.6] | 15.9 [14.9-16.9] | 16.4 [15.3-17.6] | 13.5 [11.6-15.8] |
| 55-64 | 20.6 [17.7-23.5] | 14.3 [13.3-15.2] | 14.2 [13.1-15.2] | 14.7 [12.7-17.0] |
| ≥65 | 21.8 [18.1-25.5] | 9.2 [8.4-10.0] | 9.0 [8.1-9.9] | 10.0 [8.3-11.9] |
| Missing, *n* | 0 [0-0] | 4 | 4 | 0 |
| Occupational social grade | | | | |
| ABC1 (more advantaged) | 19.3 [17.9-20.7] | 58.0 [56.6-59.3] | 58.5 [57.0-60.0] | 56.0 [53.0-59.0] |
| C2DE (less advantaged) | 20.9 [19.2-22.7] | 42.0 [40.7-43.4] | 41.5 [40.0-43.0] | 44.0 [41.0-47.0] |
| Missing, *n* | - | 0 | 0 | 0 |
| Smoking status | | | | |
| Never | 19.5 [16.4-22.7] | 11.7 [10.8-12.6] | 11.7 [10.8-12.8] | 11.4 [9.6-13.5] |
| Former | 14.1 [12.7-15.5] | 47.0 [45.6-48.3] | 50.5 [48.9-52.0] | 33.0 [30.2-36.0] |
| Current | 26.9 [25.0-28.8] | 41.3 [40.0-42.7] | 37.8 [36.3-39.3] | 55.6 [52.5-58.6] |
| Missing, *n* | - | 0 | 0 | 0 |
| Frequency of smoking[4] | | | | |
| Daily | 30.2 [27.7-32.7] | 67.9 [65.8-70.0] | 65.3 [62.7-67.7] | 75.0 [71.1-78.5] |
| Non-daily | 21.3 [18.1-24.5] | 32.1 [30.0-34.2] | 34.7 [32.3-37.3] | 25.0 [21.5-28.9] |
| Missing, *n* | - | 0 | 0 | 0 |

[1] Among eligible participants – i.e., those who reported current vaping and no concurrent use of nicotine replacement therapy, heated tobacco, or nicotine pouches. The proportions with missing data include those who responded that they did not know and those who reported zero expenditure.
[2] Eligible participants who reported their weekly expenditure on vaping products.
[3] Eligible participants did not report their weekly expenditure on vaping products (i.e., responded that they did not know or reported zero expenditure).
[4] Among those who also smoke cigarettes.

**Table S2.** Distribution of weekly inflation-adjusted expenditure (in £) on vaping products among people (≥16y) in England who vape

| | Column % [95% CI][1] | | | | |
|---|---|---|---|---|---|
| **Weekly expenditure (£)** | **2021** | **2022** | **2023** | **2024** | **2025** |
| <5 | 30.7 [27.9–33.7] | 21.8 [18.7–25.2] | 21.7 [18.9–24.8] | 17.6 [14.6–21.0] | 16.1 [12.4–20.6] |
| 5 – <10 | 32.2 [29.3–35.2] | 27.7 [24.2–31.6] | 25.3 [22.2–28.6] | 24.8 [21.5–28.5] | 31.3 [26.3–36.8] |
| 10 – <15 | 21.5 [19.0–24.2] | 26.0 [22.6–29.6] | 22.9 [19.9–26.2] | 28.7 [25.1–32.7] | 26.4 [21.7–31.7] |
| 15 – <20 | 5.9 [4.6–7.5] | 7.6 [5.8–9.9] | 8.8 [6.9–11.1] | 7.3 [5.4–9.7] | 10.4 [7.4–14.6] |
| ≥20 | 9.8 [8.0–11.9] | 16.9 [14.0–20.4] | 21.3 [18.4–24.6] | 21.6 [18.3–25.2] | 15.8 [12.2–20.2] |

CI, confidence interval.

[1] Data are weighted proportions aggregated across participants surveyed within each year.

**Table S3.** Unmodelled annual estimates of weekly inflation-adjusted expenditure (in £) on vaping products among people (≥16y) in England who vape

| | Mean [95% CI] expenditure (£) | | | | |
|---|---|---|---|---|---|
| | **2021** | **2022** | **2023** | **2024** | **2025** |
| Overall | 6.79 [6.41–7.19] | 8.54 [7.95–9.19] | 8.76 [8.20–9.36] | 9.09 [8.45–9.78] | 8.26 [7.58–8.99] |
| Main device type | | | | | |
| Reusable | 6.64 [6.25–7.05] | 7.51 [6.89–8.19] | 7.23 [6.63–7.88] | 7.79 [7.20–8.43] | 7.50 [6.79–8.27] |
| Disposable | 9.36 [7.37–11.89] | 11.35 [9.96–12.94] | 11.39 [10.34–12.55] | 12.92 [11.16–14.97] | 10.22 [8.69–12.04] |
| Age (years) | | | | | |
| 16-24 | 8.14 [6.97–9.50] | 10.30 [8.97–11.83] | 11.63 [10.23–13.22] | 12.65 [11.02–14.52] | 8.98 [7.59–10.64] |
| 25-34 | 7.29 [6.49–8.18] | 9.79 [8.57–11.19] | 10.45 [9.15–11.94] | 9.50 [8.17–11.03] | 7.78 [6.74–8.99] |
| 35-44 | 8.17 [7.18–9.30] | 7.34 [6.09–8.86] | 9.95 [8.79–11.25] | 8.78 [7.34–10.51] | 9.23 [7.25–11.75] |
| 45-54 | 6.17 [5.42–7.04] | 8.01 [6.68–9.60] | 5.68 [4.84–6.67] | 8.14 [7.09–9.33] | 8.78 [6.95–11.09] |
| 55-64 | 5.31 [4.75–5.94] | 5.94 [4.96–7.11] | 5.97 [4.89–7.29] | 6.24 [5.26–7.40] | 7.66 [6.02–9.75] |
| ≥65 | 4.65 [3.84–5.63] | 5.74 [4.32–7.62] | 5.01 [4.08–6.16] | 5.39 [3.93–7.39] | 5.81 [4.23–7.98] |
| Occupational social grade | | | | | |
| ABC1 (more advantaged) | 6.87 [6.39–7.38] | 8.87 [8.07–9.75] | 8.39 [7.69–9.14] | 9.83 [9.02–10.70] | 8.35 [7.47–9.33] |
| C2DE (less advantaged) | 6.73 [6.18–7.33] | 8.28 [7.44–9.22] | 9.11 [8.26–10.05] | 8.52 [7.61–9.55] | 8.18 [7.23–9.26] |
| Smoking status | | | | | |
| Never | 5.51 [4.48–6.77] | 8.83 [7.11–10.96] | 9.98 [8.47–11.77] | 9.82 [8.21–11.73] | 8.31 [6.93–9.95] |
| Former | 6.79 [6.32–7.31] | 7.58 [6.86–8.37] | 8.34 [7.55–9.21] | 8.71 [7.94–9.55] | 8.09 [7.26–9.01] |
| Current | 7.01 [6.37–7.72] | 9.55 [8.50–10.73] | 8.81 [7.95–9.77] | 9.36 [8.04–10.88] | 8.53 [7.11–10.22] |
| Frequency of smoking[2] | | | | | |
| Daily | 6.94 [6.13–7.85] | 8.81 [7.52–10.32] | 7.67 [6.66–8.84] | 9.72 [7.68–12.30] | 6.52 [4.84–8.77] |
| Non-daily | 6.97 [5.89–8.25] | 9.80 [8.12–11.83] | 11.05 [9.46–12.92] | 9.49 [7.68–11.74] | 10.54 [8.47–13.11] |

CI, confidence interval.

[1] Data are weighted estimates of geometric mean expenditure aggregated across participants surveyed within each year.

[2] Among those who also smoke cigarettes.

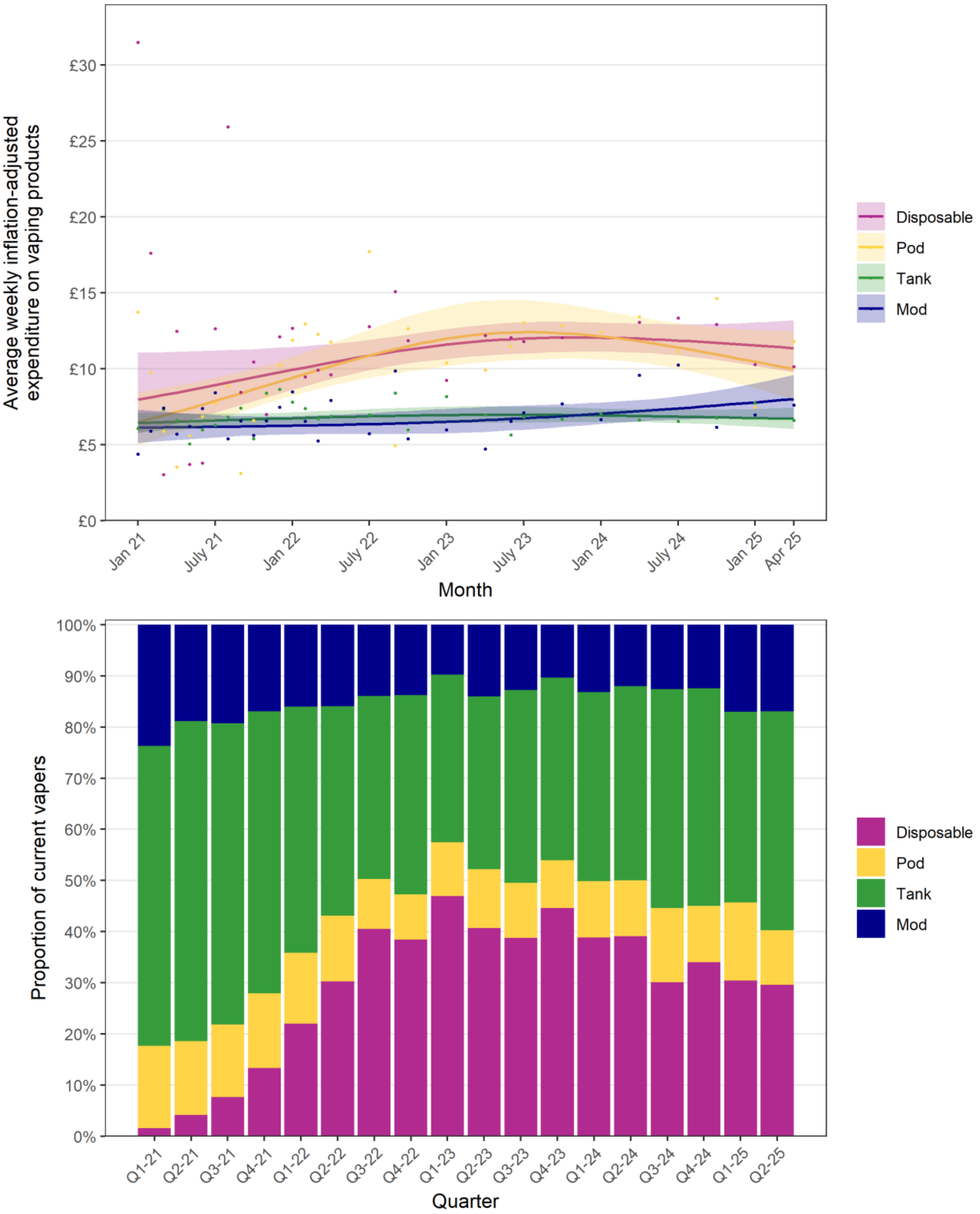


**Figure S1. Trends in expenditure on vaping products and proportions using different device types among people in England who vape, January 2021 to April 2025.** The top panel shows the modelled weighted (geometric) mean inflation-adjusted weekly expenditure (in £) on vaping products by monthly survey wave, modelled non-linearly using restricted cubic splines (three knots), by the main device type used. Lines represent mean values, shaded bands represent 95% confidence intervals, and points represent unmodelled monthly data points. The bottom panel shows the unmodelled (observed) proportion of adults who vape mainly/exclusively using each device type by quarter.
